# Automated Deep Learning-Based 3D-to-2D Segmentation of Geographic Atrophy in Optical Coherence Tomography Data

**DOI:** 10.1101/2025.07.07.25330902

**Authors:** Hasenin Al-khersan, Jonathan Oakley, Daniel Russakoff, Jessica Cao, Stanley Saju, Avery Zhou, Simrat Sodhi, Niveditha Pattathil, Netan Choudhry, David Boyer, Charles Wykoff

**Author notes:** **Corresponding author** Jonathan D. Oakley.

## Abstract

**Purpose:** We report on a deep learning-based approach to the segmentation of geographic atrophy (GA) in patients with advanced age-related macular degeneration (AMD).

**Method:** Three-dimensional (3D) optical coherence tomography (OCT) data was collected from two instruments at two different retina practices. This totaled 367 and 348 volumes, respectively, of routinely collected clinical data. For all data, the accuracy of a 3D-to-2D segmentation model was assessed relative to ground-truth manual labeling.

**Results:** Dice Similarity Scores (DSC) averaged 0.824 and 0.826 for each data set. Correlations (r^2^) between manual and automated areas were 0.883 and 0.906, respectively. The inclusion of near Infra-red imagery as an additional information channel to the algorithm did not notably improve performance.

**Conclusion:** Accurate assessment of GA in real-world clinical OCT data can be achieved using deep learning. In the advent of therapeutics to slow the rate of GA progression, reliable, automated assessment is a clinical objective and this work validates one such method.

## Introduction

Geographic atrophy (GA) is a late-stage manifestation of non-exudative age-related macular degeneration (AMD), representing one of the leading causes of vision loss in adults over 50 years of age. GA is characterized by the progressive degeneration of the retinal pigment epithelium (RPE) and overlying photoreceptors, resulting in irreversible central vision loss [Sarda 2021].

Recently, two intravitreal anti-complement therapies, pegcetacoplan and avacincaptad pegol, were introduced in the United States for the treatment of GA. These therapies were approved by the Food and Drug Administration based on structural endpoints. Both therapies demonstrated a slower rate of progression of GA relative to sham though neither showed a visual acuity benefit [Heier 2023]. Given the introduction of these therapies, the identification of GA and its progression is of heightened importance.

GA is typically diagnosed using non-invasive imaging techniques including optical coherence tomography (OCT), fundus autofluorescence (FAF), and fundus photography. OCT is particularly useful for identifying GA due to its widespread availability in clinical practice, ability to identify subtle morphological changes in early disease, and ease of capture. In 2018, OCT was proposed as the gold standard to diagnose GA by the international expert Consensus Definition for Atrophy Associated with Age-Related Macular Degeneration on OCT group [Sadda 2018]. Specific OCT criteria to diagnose complete RPE and outer retinal atrophy (cRORA) were proposed. While these criteria may accurately define GA, they are challenging for ophthalmologists to methodically apply across hundreds to thousands of B-scans in routine clinical practice. Toward this end, artificial intelligence (AI) can serve to simplify the process by automating the identification of GA.

Different approaches to segmenting GA in image data have been developed by both device manufacturers and research groups. For OCT, the only software with clinical approval in the US is the advanced retinal pigment epithelial (RPE) analysis tool (ARPET; Carl Zeiss Meditec, CA, USA). Marketed now as a GA tool kit, it was approved to automatically segment sub-RPE “increased illumination” based on an en face view generated using a layer-based limited integration range [K111157 2012]. The success of any 2D approach based on a limited axial integration of the 3D volume is entirely dependent on the relevance and location of the automatically generated integration limits. The approval of this method for clinical use was based on repeatability (precision) and, interestingly, correlation to the sum areas of manually segmented GA in FAF images, but not to the location accuracy of those areas as could be reported using the Dice similarity coefficient (DSC). This is the same in a study of the algorithm, where the intraclass correlation coefficient (ICC) was 0.795, but no accuracy of segmentation localization was reported [Yehoshua 2013].

The advent of deep learning has resulted in several fully automated applications. Available via the Heidelberg AppWay product, users in the EU may access the GA model developed by RetinAI (Bern, Switzerland). The method is based on segmenting each B-scan using a U-Net individually to yield a 2D probability image of the atrophic region [Derradji 2021]. This result is then integrated axially to form a 1d signal that, when combined with all other B-scan-based 1d signals, reconstructs the final 2D image of GA. Using a training set of 44 volumes, the authors are able to demonstrate high DSCs of 0.881 and 0.844 relative to two graders on a test set of 18 volumes. The method’s advantage is its simplicity; its disadvantage is it not making full use of the 3D data set, which can result in discontinuities in the final segmentation as each B-scan is segmented independently. A very similar approach has been used by [Zhang 2021], but a far larger data set of 984 OCT volumes was used, and the *median* DCS was very high at 0.96.

[Lachinov 2021] overcomes some limitations of the aforementioned approaches by working directly with the 3D data using a then novel architecture that modifies a U-Net to encode in 3D and decode in 2D. In this approach, the skip connections, common to the U-Net architecture, drop one dimension from the data and go from 3D to 2D. Our extension to that method adds residual blocks to allow for better feature representations [He 2016]. In the following we examine the ability of this 3D U-Net-type architecture to identify GA on OCT data from real-world patient data.

## Methods

### Data Collection and Grading

Data was collected from two sites retrospectively, the Retina Consultants of Texas (RCTX) and Retina-Vitreous Associates (RVA), from 2016 and 2022. Inclusion criteria included a confirmed diagnosis of GA with accompanying OCT imaging; eyes with GA and concurrent nAMD were included. Institutional review board approval was obtained from Advarra. The present study was conducted in accordance with the Declaration of Helsinki.

Optical coherence tomography imaging was performed on the Heidelberg Spectralis (Heidelberg Engineering, Heidelberg, Germany) at RCTX and the Zeiss Cirrus (Carl Zeiss Meditec, CA, USA) at RVA as part of routine clinical care. Both used similar protocols for capturing data at the macula using a 20 degree (∼6mm^2) lateral field of view (FOV).

For Spectralis, the macular scan protocol was used with ART setting of up to 10 averaged frames. The volumes comprised 49 slices, where each B-scan was 512 by 496 pixels. The Spectralis data included near-infrared (nIR) images, which were across a 30-degree FOV and used 100 frame averaging. A quality index of at least 25 was required for inclusion.

For Cirrus data, the macular protocol included 200 B-scans, each with 200 by 1024 pixels. The Zeiss macular protocol does not use frame averaging. A minimal signal score of 5 was required for inclusion. No associated nIR data was collected by the device.

#### Grading

Manual grading was performed by expert graders who delineated areas of complete retinal pigment epithelial and outer retinal atrophy (cRORA) on OCT based on the Classification of Atrophy Meetings (CAM) criteria as previously described.^1^ Graders marked areas of cRORA using the Orion (Voxeleron Inc., TX, USA) application [Alex 2021]. Specifically, graders marked the bounds of atrophy on the B-scan cross-sections (Figure 1). The delineated areas are subsequently represented two-dimensionally on the OCT en face view for confirmation of correct demarcation. Graders were masked to the nIR images collected in the Spectralis data set for consistency with the Cirrus labeling, which had no nIR data.

**Figure 1.**
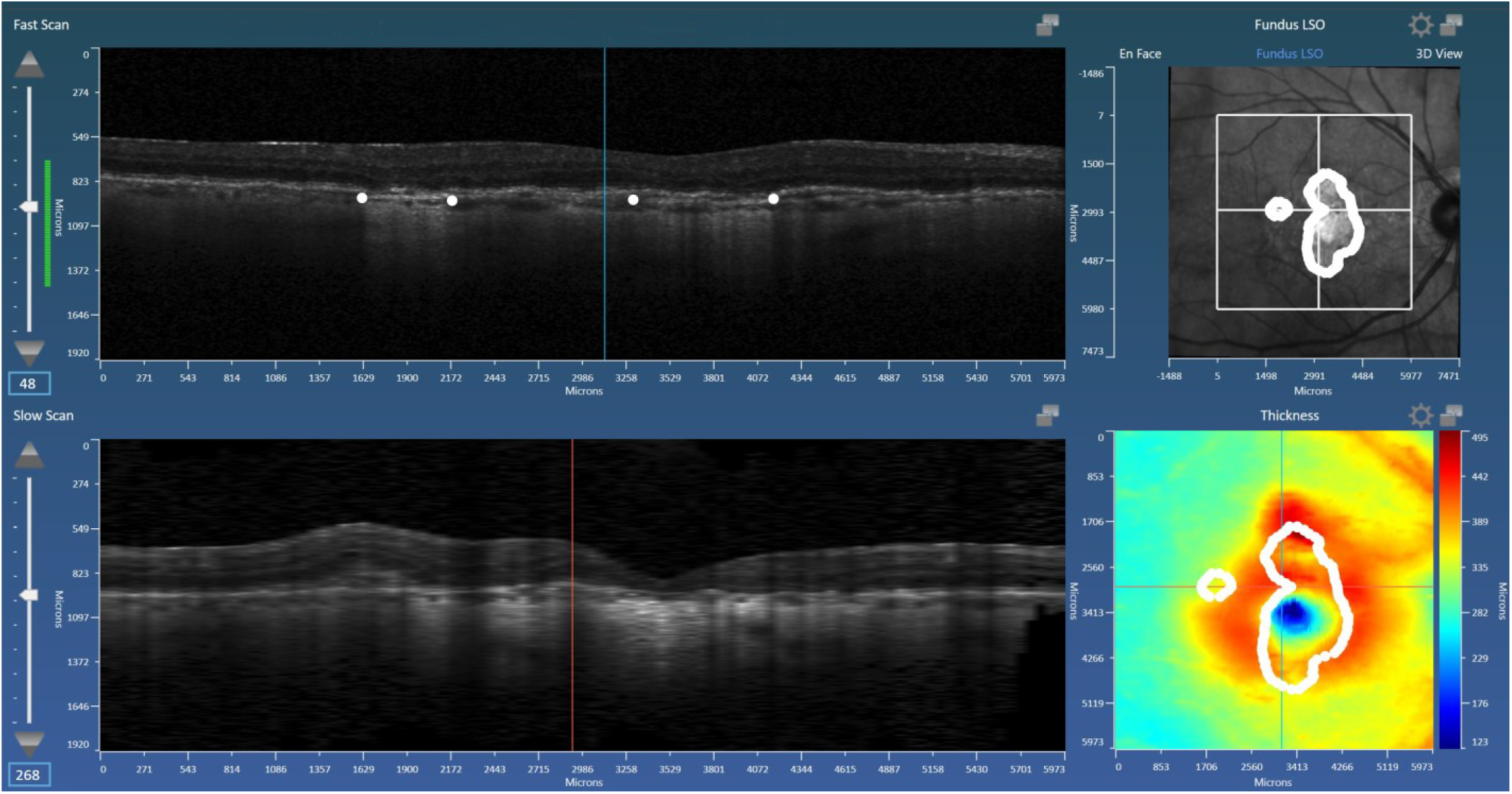
The graders used Orion to place landmarks at each of the end points of the atrophy in each B-scan. The resulting 2D mask (area) is shown in the OCT en face view (top right). Once the editing is complete, the results file is saved and the mask exported as our ground truth area delineation.

### Deep Learning Architecture

A 3D U-Net architecture was used for the deep learning with the output modified to generate a 2D mask. It extends the approach of [Lachinov 2021] by adding attention blocks to the architecture in an effort to better localize the region in which the atrophy is imaged. Explicit localization is done in both [Yehoshua 2013] and [Derradji 2021] using layer segmentation, but here we do this in 3D and in a single architecture with the localization learned. A schematic of the 3D-to-2D U-Net is given in Figure 2.

**Figure 2.**
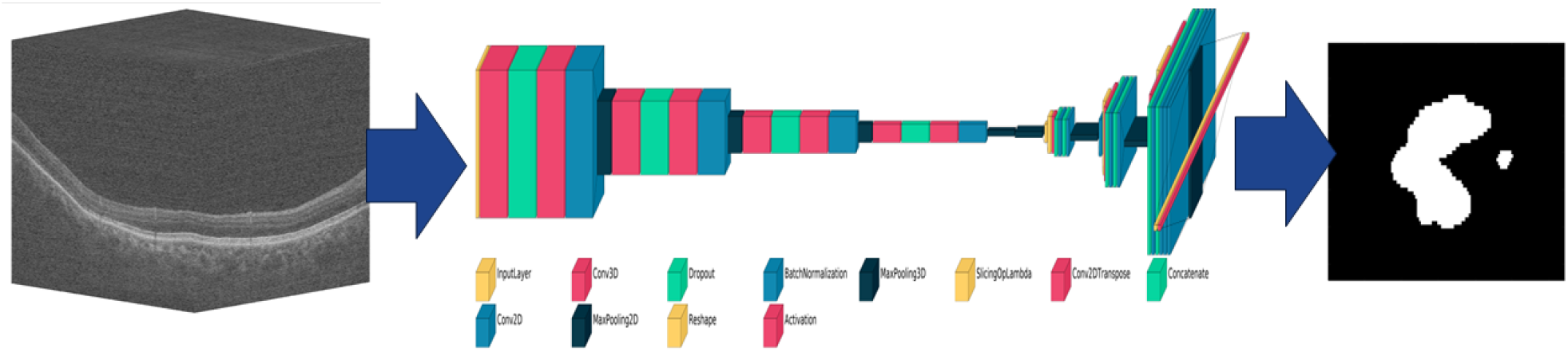
Schematic of the CNN-based 3D to 2D neural network used for this study.

To alleviate memory management issues caused by large 3D volumes, in the Spectralis data the volume was sub-sampled to 128-by-128-by-64 pixels (i.e., to 64 B-scans each of 128^2 pixels). The Cirrus data, due to the more isotropic distribution, was sub-sampled instead to 128-by-128-by-128 pixels (i.e., to 128 B-scans each of 128^2 pixels). The output was a 2D mask of GA at a resolution of 128-by-64 pixels and 128-by-128 pixels for Spectralis and Cirrus, respectively. For the Spectralis implementation, the architecture was also capable of receiving the 2D nIR images in a second input channel to see if this additional information would improve the algorithm’s performance.

The implementation used TensorFlow 2.15 running under a Linux system on a Dell Precision 7920 Workstation, housing two NVIDIA A6000s connected via the NVLINK system allowing for distributed learning and shared GPU memory.

The parameters only differed in batch size as the Cirrus volumes were larger. The optimization algorithm used was AdamW, with a learning rate of 5e-5. The loss function used the DSC, with even weighting of 0.5 to each class (GA, non-GA). Training each of the models used 450 epochs with a batch size of 32 for Spectralis, and 20 for Cirrus data. A patience parameter was set to 70 meaning that, if after 70 iterations no improvement was seen in the performance relative to the validation split (a part of the training set), the learning stopped.

### Analysis Methods

N-fold cross-validation was performed. In this scenario, data is split N times, where at each of the N-folds, 1/Nth of the entire data set is set aside for testing and the remaining for training. Training further splits the data into train and validate groups, where the latter is used to determine when the training iterations stop. For the present study, five folds were used. For repeat cases from the same subject, the folds were structured such that at no time was data from a single subject represented both in the train and test sets. Dice Similarity Scores (DSC) and correlation (r^2^) between manual and automated areas of atrophy were calculated. All final analysis was done using Matlab and its Statistics Toolbox (Mathworks, Natick, MA, USA).

## Results

For the Spectralis data, 367 scans were used from 55 subject eyes and 33 subjects. The data was collected over a period of eight years. All 367 scans had confirmed GA: 267 demonstrated concurrent nAMD while the remaining 100 had GA without nAMD. The Cirrus data subset was comprised of 348 subject eyes from 326 subjects representing 348 total scans. All cases had confirmed GA, but 101 were also undergoing active treatment for fluid.

For the Spectralis data, the mean DSC score was 0.826 and r^2^ was 0.906 (Table 1 – Summary analysis results for all data and experiments.). There was little change in the mean DSC score (0.829) and r^2^ (0.908) with the addition of nIR data. For the Cirrus data, the mean DSC score was 0.824 and r^2^ was 0.883. Splitting the Spectralis results into GA only and GA and concurrent nAMD, we interestingly see an increase in DSC scores where nAMD is present, but far better area correlations in GA only cases. In the case of the average correlation scores, the difference was notable in the Spectralis data; for the DSC scores the changes were only slight.

**Table 1.** Summary analysis results for all data and experiments.

|  | **Cirrus** |  | **Spectralis OCT only** |  | **Spectralis OCT + nIR** |  |
| --- | --- | --- | --- | --- | --- | --- |
|  | GA only | GA + Treatment | GA only | GA + nAMD | GA only | GA + nAMD |
|  | N=247 | N=101 | N=100 | N=267 | N=100 | N=267 |
| **Mean DSC** | 0.824 |  | 0.826 |  | 0.829 |  |
|  | 0.816 | 0.845 | 0.803 | 0.834 | 0.825 | 0.830 |
| **Correlation ($R^2$)** | 0.883 |  | 0.906 |  | 0.908 |  |
|  | 0.887 | 0.869 | 0.97 | 0.85 | 0.983 | 0.845 |
|  | GA only | GA + Treatment | GA only | GA + nAMD | GA only | GA + nAMD |
|  | N=247 | N=101 | N=100 | N=267 | N=100 | N=267 |

Figure 3 shows the scatter plot and Bland-Altman plot for automated and manual measurements. Figures 4 and 5 show the same but for the Spectralis using just the OCT data, and using OCT + nIR data, respectively. Figures 6 through 9 show example segmentations for both the Spectralis and Cirrus data.

**Figure 3.**
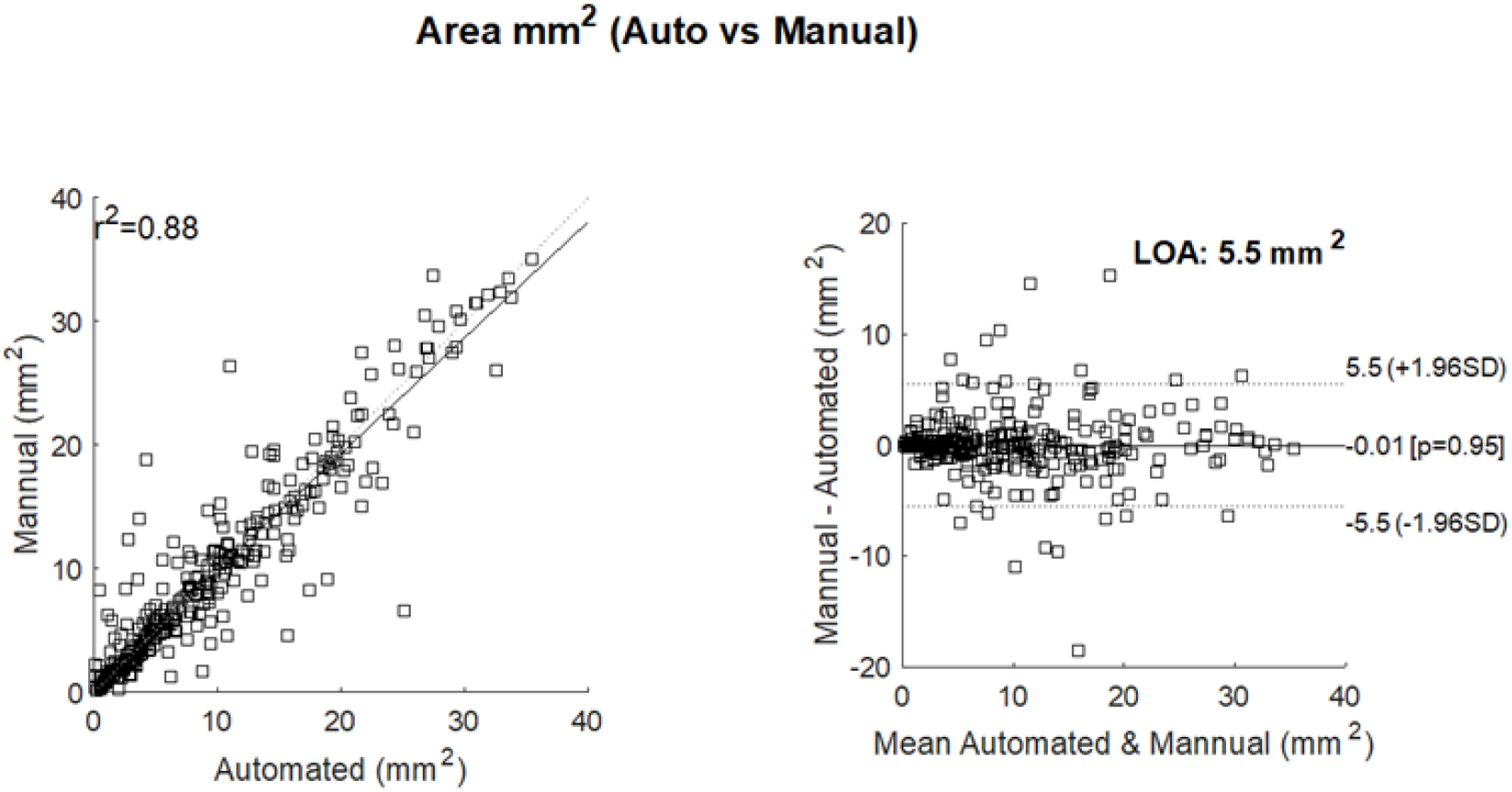
Correlation of area measurements for the Cirrus data set. Left: scatter plot showing the manual area measurements (y axis) versus the automated measurements (x-axis) and their resulting correlation (r^2^ = 0.88). Right: Bland-Altman plot showing the limits of agreement (LOA) between automated and manual measurements. The y axis shows the difference between the manual and automated measurements; the x-axis their average.

**Figure 4.**
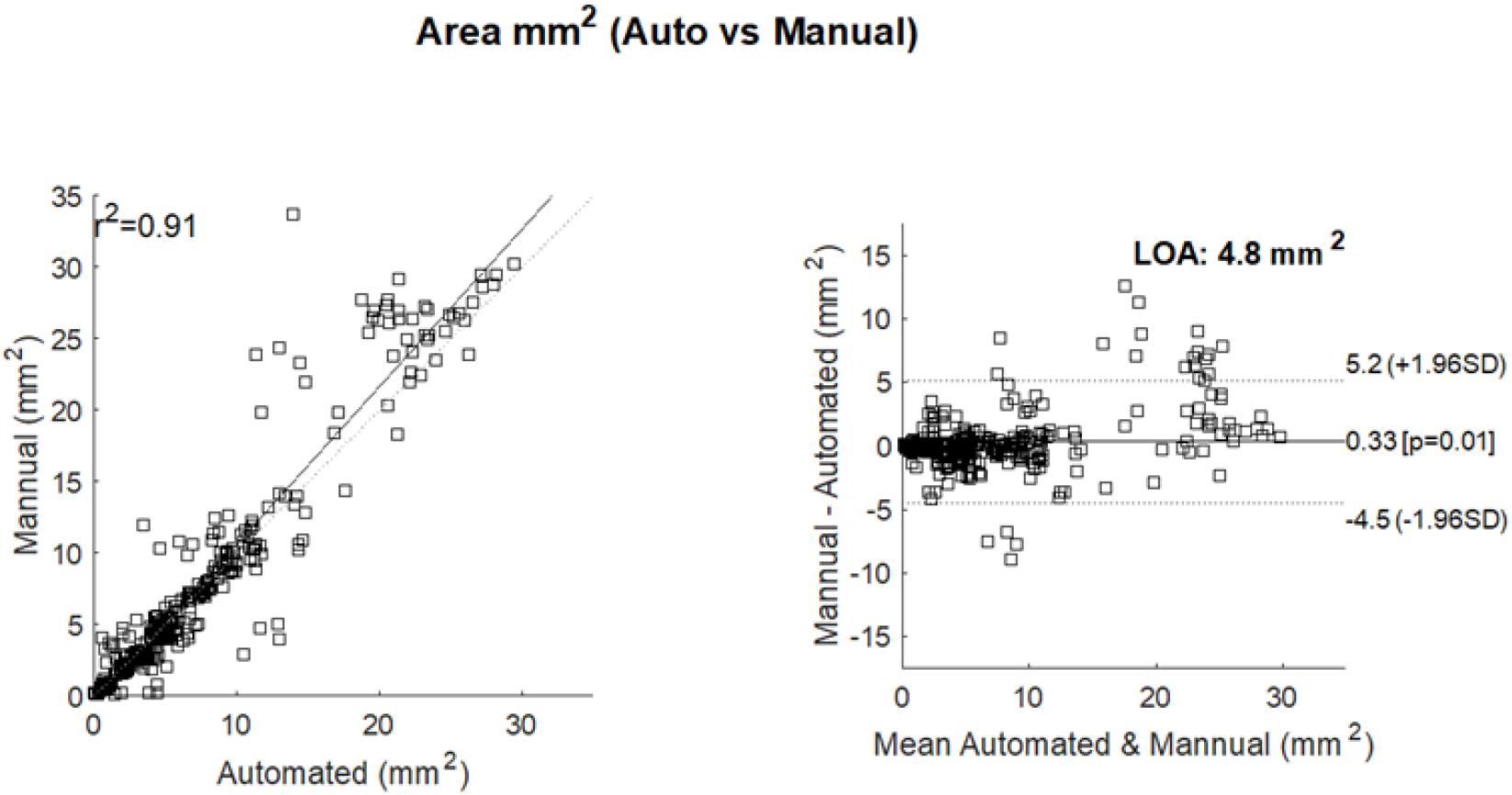
Correlation of area measurements for the Spectralis data set. Left: scatter plot showing the manual area measurements (y axis) versus the automated measurements (x-axis) and their resulting correlation (r^2^ = 0.92). Right: Bland-Altman plot showing the limits of agreement (LOA) between automated and manual measurements. The y axis shows the difference between the manual and automated measurements; the x-axis their average.

**Figure 5.**
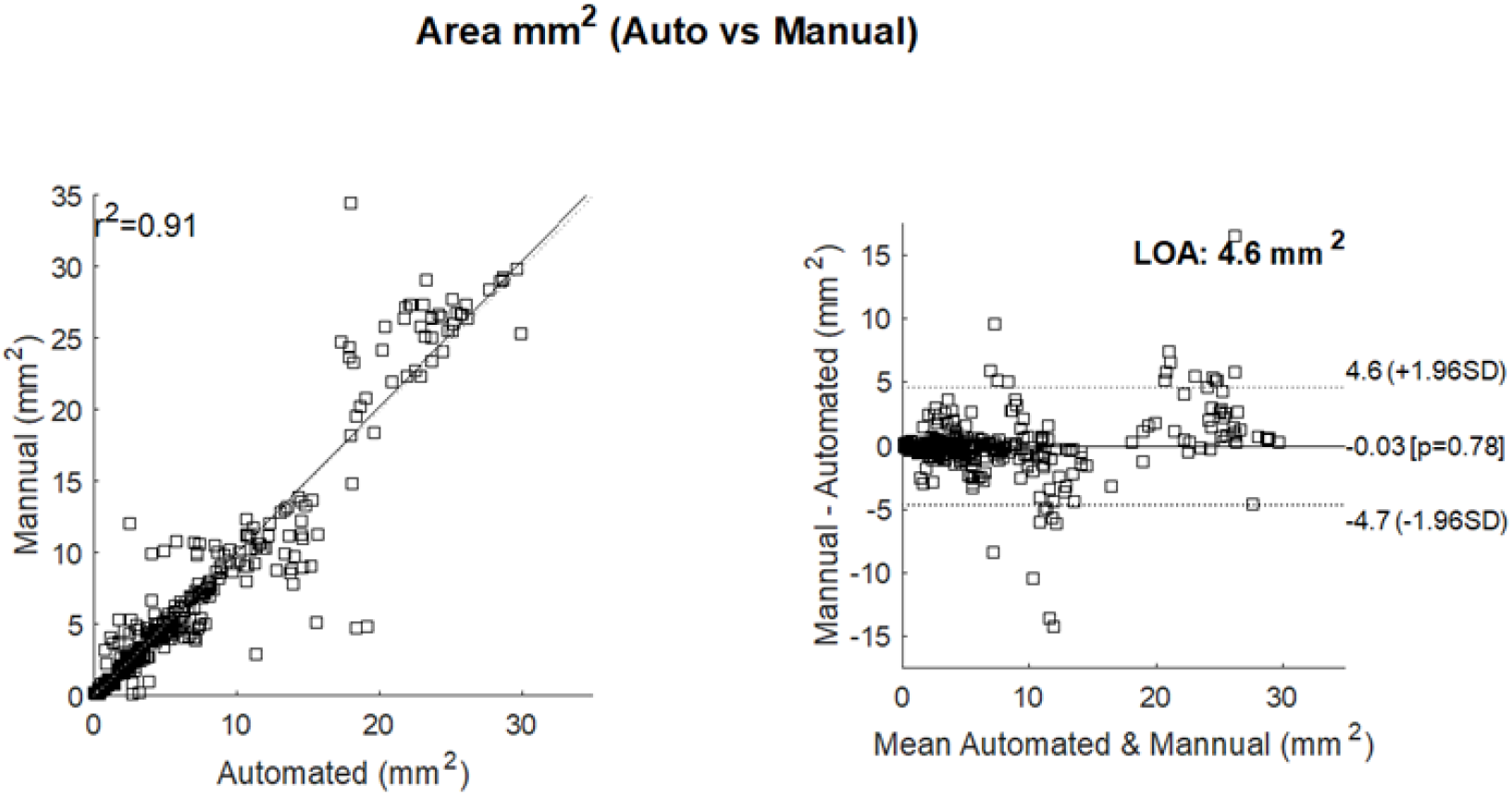
Correlation of area measurements for the Spectralis OCT + nIR data set. Left: scatter plot showing the manual area measurements (y axis) versus the automated measurements (x-axis) and their resulting correlation (r^2^ = 0.91). Right: Bland-Altman plot showing the limits of agreement (LOA) between automated and manual measurements. The y axis shows the difference between the manual and automated measurements; the x-axis their average.

**Figure 6.**
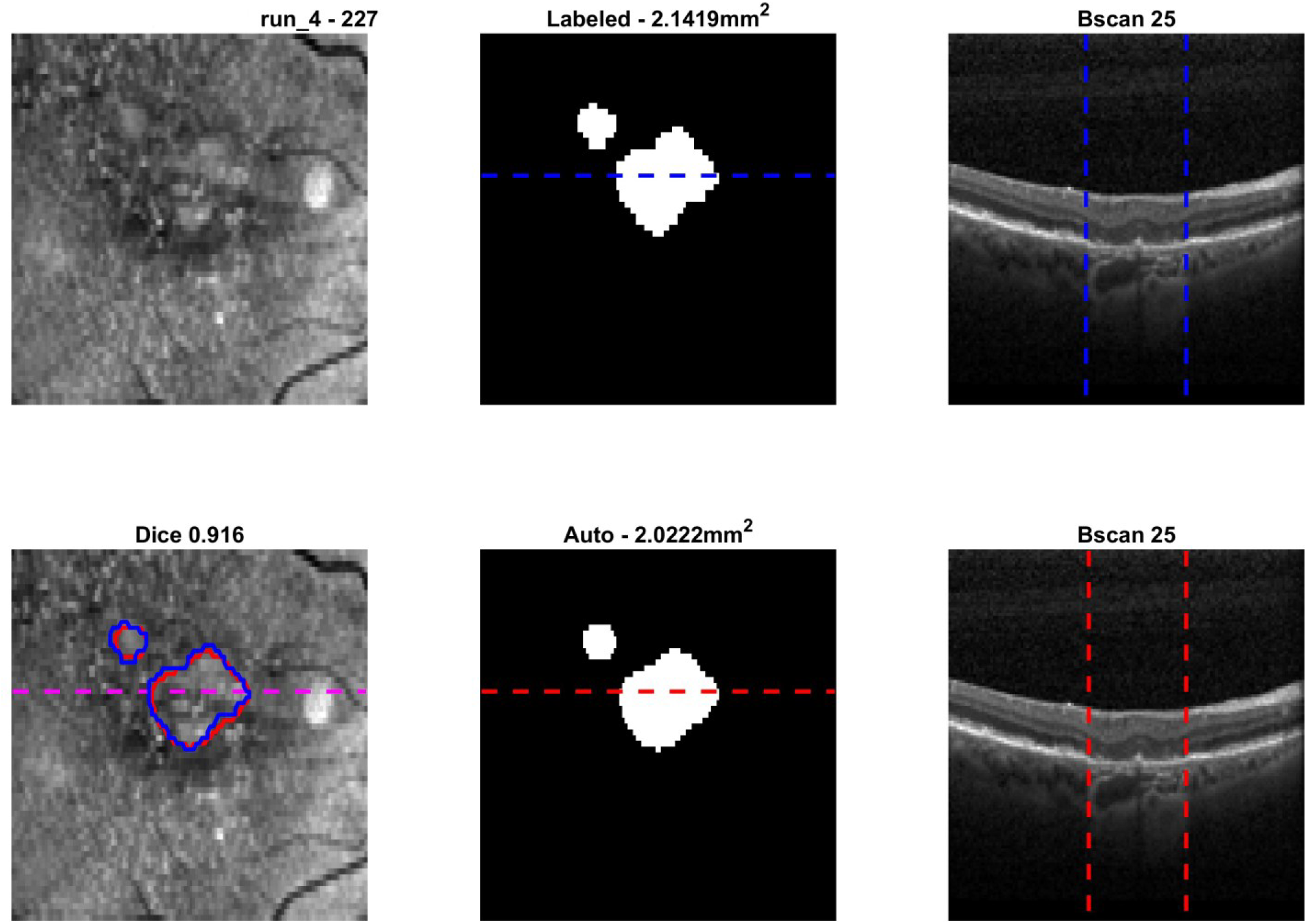
Example Spectralis data. Clockwise from top left we have: input nIR en face image; manual area delineation (blue) and automated area delineation (red); example B-scan showing the manual atrophy delineation; example B-scan showing the automated delineation; final automated mask and dashed line showing the location of the presented B-scan; final manual mask and a dashed line showing the location of its presented B-scan.

## Discussion

The FDA has recognized the area of geographic atrophy as a primary endpoint in the development of treatments for GA. This important designation means that GA area, as measured using imaging, is considered relevant and clinically meaningful. This designation is, however, limited to using fundus autofluorescence which works by detecting natural fluorescence emitted by lipofuscin, a pigment that accumulates in the retinal pigment epithelium (RPE). This modality, however, offers no cross-sectional information and obscures foveal involvement that can lead to misinterpretation of the extent of the GA and risk to vision. Within a clinical setting, OCT is the standard of care and, given its depth resolution, is capable of revealing outer retinal disruption that is prognostic of GA progression [Vogl 2023]. In the advent of complement inhibitor-based treatments for slowing the progression of GA is it important that clinicians have the tools to measure the endpoint of GA area automatically and thereby gauge the effectiveness of the treatments they are using.

The current work has presented a means for doing this automatic monitoring, looking at how well the automated approach compares to expertly labeled data. Importantly, we have used two widely used OCT devices from two different manufacturers, where the data has been collected from two different sites. Important also is the fact that this is real-world clinical data, and not data from a clinical trial. Relative to ground truth assessment, we have shown for both devices excellent correlation, narrow limits of agreement, and very high DSC scores. This is also true in cases of different ocular comorbidities, as was predominantly seen in the data from the Cirrus device.

The results are encouraging with respect to the presence concurrent nAMD and other comorbidities that involve fluid. This, perhaps, is due to the fact that fluid reflects very little light and does not, therefore, attenuate the signal that penetrates through to the choroid. It is the hyperreflectivity that is such a distinctive feature of cRORA in OCT volumes.

Ground truth data is, of course, subjective and while the grading was strictly based on adherence to the CAM classification’s definition of cRORA, it is difficult for graders to be consistent across large data sets of variant image quality. This, however, is a general limitation and is confirmed in the variability seen in other studies [Chandra 2022, Schmitz-Valckenberg 2023], and something that we have not investigated in this study.

The approach adopted in this study takes the entire OCT volume as input and returns a 2D area as a result. To do this we have created a quite bespoke architecture based around a residual U-Net. An alternate approach is to process each 2D B-scan separately and combine outputs to produce a final result. These approaches are based on segmenting each B-scan to yield a 2D probability image of the atrophic region [Derradji 2021, Zhang 2021]. This result is then integrated axially to form a 1d signal that, when combined with all other B-scan-based 1d signals, reconstructs the final 2D image of GA. This has the advantage of having a low overhead in terms of memory requirements, but the disadvantage in losing the context of the 3D data. Subsequently, this can result in segmentation areas having discontinuities between slices.

While solving for this, the 3D data processing we have chosen incurs significant overhead in terms of memory requirements for model building. Our solution has been to reduce the size of the volumes by sub-sampling. But while this can have a nice denoising effect, the fidelity of the data comes into question, meaning detail may be lost, which is possibly the case in Figure 7. This can be circumvented through better memory management or indeed larger GPUs, and can be immediately solved by using large server clusters. However, these solutions come at a high financial cost. Interestingly though, where the data was more isotropic, that is the Cirrus used 128 B-scans as opposed to the Spectralis’ 64 B-scans, this issue goes away (Figure 8). For 3D processing, isotropic data is far more preferrable. The Spectralis protocol collects at least as much data, but gives preference to averaging often sparsely spaced B-scans as opposed to creating a dense, but unaveraged cube.

**Figure 7.**
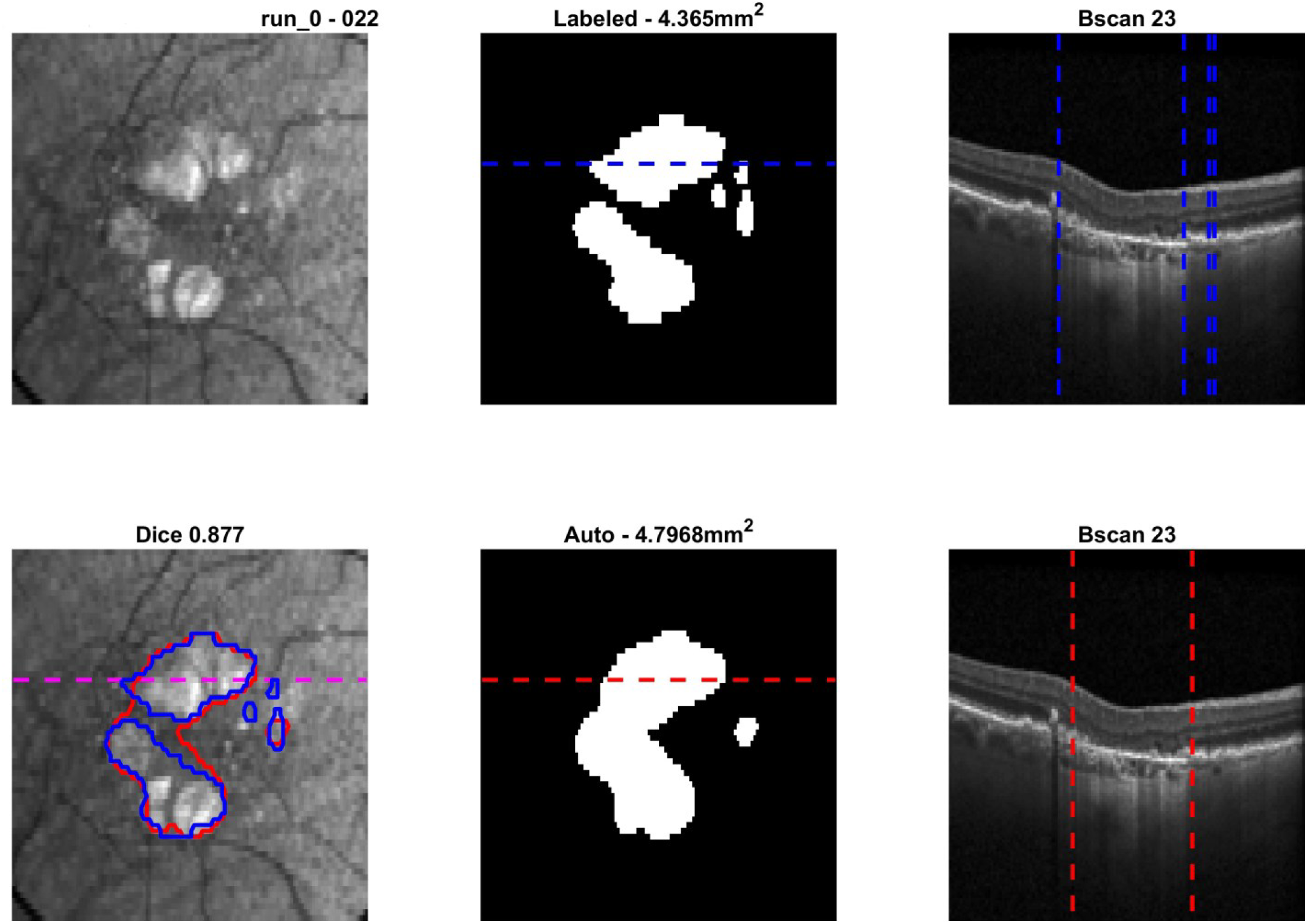
Example Spectralis data. Clockwise from top left we have: input nIR en face image; manual area delineation (blue) and automated area delineation (red); example B-scan showing the manual atrophy delineation; example B-scan showing the automated delineation; final automated mask and dashed line showing the location of the presented B-scan; final manual mask and a dashed line showing the location of its presented B-scan.

**Figure 8.**
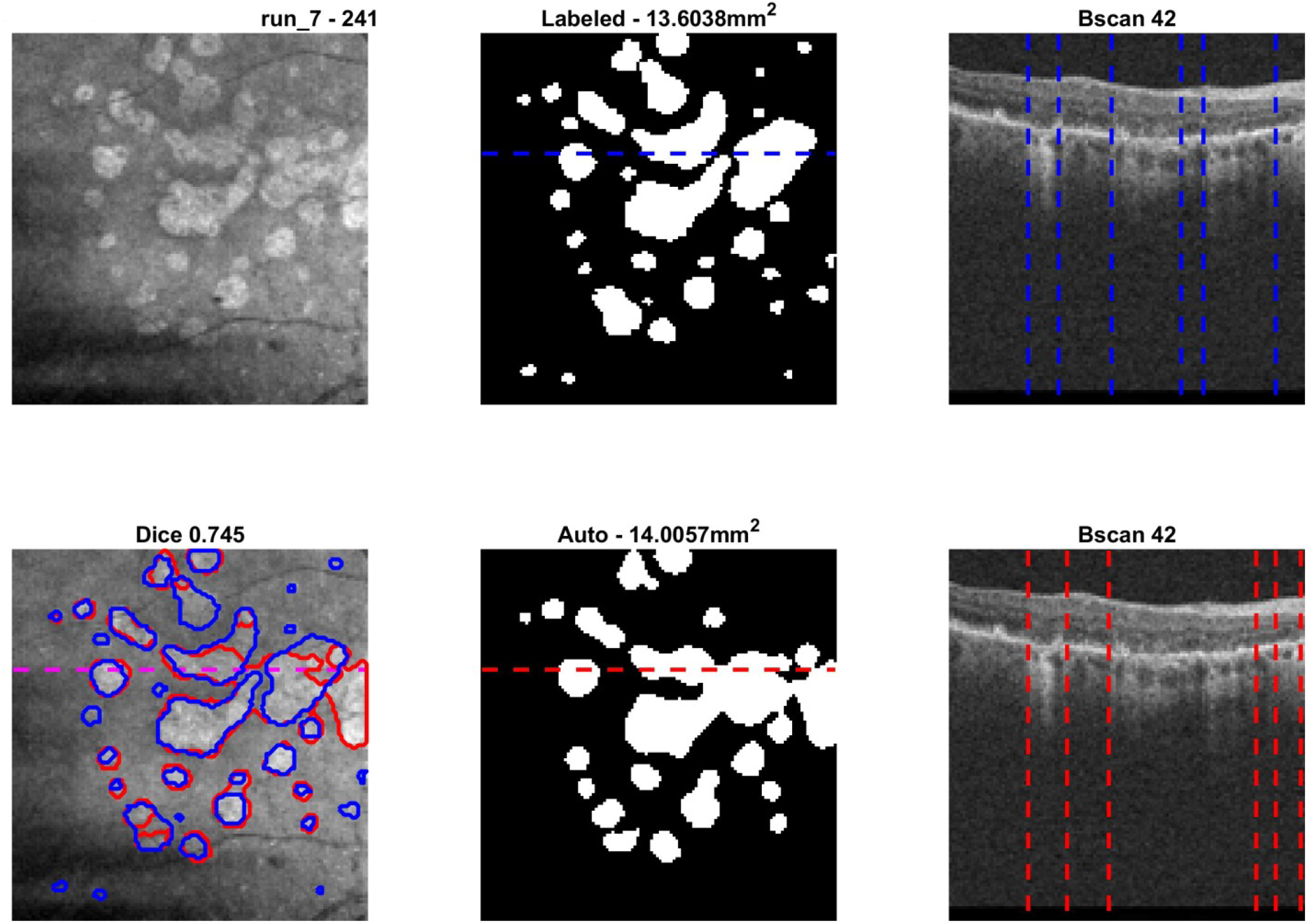
Example Cirrus data. Clockwise from top left we have: input OCT en face image; manual area delineation (blue) and automated area delineation (red); example B-scan showing the manual atrophy delineation; example B-scan showing the automated delineation; final automated mask and dashed line showing the location of the presented B-scan; final manual mask and a dashed line showing the location of its presented B-scan.

Our study is not, however, without some limitations that must be considered in interpreting these results.

For the Spectralis grading it was a design decision not to use the nIR images during the grading. Given this image data is quite unique to that device, this would go against the ultimate objectives of having a device agnostic automated approach. This, however, will reduce a grader’s ability to accurately assess GA as additional modalities are complementary and supportive of a final assessment. This leads to the question of accuracy of the ground truth data when not all information was available. Indeed, we have seen in the work of [Lachinov et al.], who used FAF in their grading, and just OCT in their segmentations, their performance dropped. It makes intuitive sense that the labeling task, the visual task, must use the same modality as that given to the segmentation algorithm. The main visual cues for cRORA are the absence of RPE and hyper transmission defects. It is possible that the network only learns the latter, but this might not be a limitation given that the appearance and growth of persistent choroidal hypertransmission defects in eyes with iAMD is an early sign of GA, making it a candidate for clinical trial endpoints [Liu2023]. And iRORA, cRORA both require hypertransmission defects and do not require a completely absent RPE.

Using the OCT data alone likely has the consequence of a potential performance ceiling to the performance of the CNN. This was somewhat tested when nIR data was also given as input information to the network. In this case, we did not expect, therefore, that the CNN would improve its performance. If, however, we had graded with the user of nIR images, we might expect this to be different. It is beyond the scope of this current study to assess such permutations that incorporate different modalities, but it is an important consideration in both study design and ultimately clinical deployment to determine the appropriate combinations. We have instead adhered to two important tenets in this work: firstly, that OCT is the reference standard for diagnosis and staging of GA [Sadda 2018]; and secondly that the approach presented be applicable to any OCT device.

Lastly, Spectralis data with a noise score below the manufacturers’ recommendations were excluded from this study. Not only because it causes a drop in algorithm performance, it results in data that is harder to grade, so performance targets become increasingly ambiguous. It is, however, an open question as to whether or not such data should be included in the training data given labeling inaccuracies, and one that is not addressed in this study.

## Data Availability

All results produced in the present study are available upon reasonable request to the authors.

## Notes

### Competing Interest Statement

The authors have declared no competing interest.

### Funding Statement

This study did not receive any funding.

### Author Declarations

Institutional review board approval was obtained from Advarra. The study involved retrospective analysis of anonymized imaging data collected during routine clinical care and did not require individual patient consent. The study adhered to the principles outlined in the Declaration of Helsinki.

